# The allostatic overload in pregnancy during the COVID-19 pandemic and potential effects on the health of the mother-child dyad: Study Protocol

**DOI:** 10.1101/2025.05.22.25328155

**Authors:** Noelia Benavente-González, Patricia A. Huerta, Fernando Teillier, Aldo Sepúlveda, Susan Urrea, Javiera Cheuquelao, Patricio Castro, Pablo Vergara-Barra, Enrique Guzmán-Gutiérrez, Carlos Escudero, Marcelo González-Ortiz

## Abstract

**Introduction:** Allostatic load refers to the cumulative burden of stress and life events that involve the interaction of various physiological systems at differing levels of activity. Allostatic overload occurs when environmental challenges surpass an individual’s ability to cope. The COVID-19 pandemic has subjected pregnant women to an unprecedented amount of stress and uncertainty, and this situation has been linked to higher rates of mental health disorders, especially during lockdowns and the early stages of the pandemic. Contributing factors potentially include gender-based violence, gender inequality, multidimensional poverty, barriers to healthcare, isolation, and social restrictions, among others. Perinatal mental health disorders, such as anxiety, depression, and stress, have lasting health effects on both mothers and their children and could be significant risk factors for allostatic overload. Maternal mood and anxiety disorders during pregnancy have been linked to cardiovascular diseases, particularly ischemic heart disease, years after childbirth. Additionally, there is evidence showing the cardiovascular impacts of COVID-19, including an increased expression of thrombosis biomarkers in patients infected with SARS-CoV-2.

**Aim:** To evaluate the impact of allostatic overload during pregnancy, particularly in the context of the COVID-19 pandemic, on mothers’ cardiovascular health, children’s neurodevelopment, and the mother-child relationship.

**Study design:** We propose a historical cohort study focused on mothers who were pregnant during the COVID-19 pandemic (2020–2021). We will retrospectively assess exposure to allostatic overload during the perinatal period by triangulating both quantitative and qualitative data. The outcomes for mother-child dyads will be evaluated prospectively.

**Methods:** We propose to study 32 exposed and 64 control subjects to reject the null hypothesis that the relative risk equals 1. This sample size was estimated to detect a significantly higher relative risk of ischemic heart disease. The primary outcomes will be: 1) Manifestation or higher risk of ischemic heart disease in the mothers. 2) Neurodevelopmental disorders in children. 3) Alterations in the mother and child relationship. To assess exposure to allostatic overload, we will conduct a clinimetric survey and narrative interviews to explore life experiences and identify significant stressors during the perinatal period. The potential association between allostatic overload and health later in life will be analysed using multivariable epidemiological analyses and machine learning techniques.

**Expected results:** We anticipate a higher prevalence of allostatic overload in women who experienced pregnancy during the first wave of the COVID-19 pandemic compared to the later period. It is proposed that these women may face an increased risk of hypertension, endothelial dysfunction, and ischemic heart disease due to allostatic overload. Additionally, we expect to observe changes in plasma biomarkers related to cardiovascular health and a higher risk of neurodevelopmental disorders in their children. Furthermore, increased parental stress associated with allostatic overload during pregnancy may lead to a poorer mother-child relationship.

## 1. Background of the study

### 1.1 Allostatic overload in pregnancy

Psychosomatic medicine is an interdisciplinary field that examines the interaction between biological, psychological, and social factors in maintaining the balance between health and disease (1). A key concept in this field is "allostatic load" (AL). Allostasis is the process by which organisms actively adapt to predictable and unpredictable events. When the events and context overcome the capacity to adequately maintain the allostasis, the "Allostatic Overload" (AOL) emerges. AOL represents the stress and physical strain resulting from repeated fluctuations in physiological responses and heightened activity of physiological systems in response to challenges and changes in metabolism that can lead to disease (2).

External factors, including family, economic, and social environments, significantly influence behaviours that promote or damage health. For example, during pregnancy, factors such as income inequality, increased social instability, and gender-based violence can contribute to AOL (3). AOL is recognized as a strong indicator of chronic stress exposure and is predictive of morbidity and mortality, closely associated with conditions such as hypertension and cardiovascular disease (CVD)(4).

AOL is identified using biomarkers and clinical criteria (5), but clinical criteria should take precedence in certain situations, such as pregnancy and retrospective exposure. Pregnancy triggers a cascade of biological changes that can influence the levels of AOL-related biomarkers, including those related to cardiovascular health, inflammation, and metabolism. Therefore, measuring these biomarkers during pregnancy can be problematic (4). The evaluation of AOL in pregnant women should rely primarily on clinical criteria, considering various stressors, such as hypertensive pregnancy disorders, maternal obesity, diabetes, psychosocial stress, and prenatal anxiety or depression. Other uncontrolled events that may significantly alter the patient’s living conditions, social and family dynamics, and work situation can be assessed using qualitative methods (3).

AOL during pregnancy has been linked to poor sleep quality, which is a well-known chronic stressor (6). It is also associated with increased odds ratio (OR) for preeclampsia and preterm birth (7). While there is a lack of studies on the long-term effects of AOL during pregnancy on women and children, one study indicated that prenatal depression raises the risk of cardiovascular disease (CVD) in women two years after giving birth (8). In this study, the adjusted hazard ratios (aHRs) within the 24 months postpartum for women with prenatal depression were as follows: 1.83 for ischemic heart disease, 1.61 for cardiomyopathy, 1.60 for arrhythmia or cardiac arrest, 1.40 for heart failure, 1.32 for hypertension, and 1.27 for cerebrovascular disease or stroke. These cardiovascular effects later in life are linked to hypertensive disorders during pregnancy, which carry a 2 to 4-fold increased risk for heart failure, stroke, and ischemic heart diseases decades after childbirth (9). When combined with preterm birth, hypertensive disorders in pregnancy can increase CVD-related deaths by 5 to 7 times (10). Despite this evidence regarding the impact of prenatal depression and hypertension on women’s cardiovascular health, the potential association between AOL during the perinatal period and future cardiovascular health has not yet been explored. This study aims to address that gap in the field of psychosomatic medicine.

### 1.2 Allostatic overload and early development

The Developmental Origins of Health and Disease (DOHaD) theory posits that perinatal stress triggers adaptive changes in endocrine and metabolic systems, which can become permanently programmed and influence health in adulthood (11). The AOL concept could be central to the DOHaD theory because the physiological mechanisms during early human development are under significant pressure. The hypothalamic-pituitary-adrenal (HPA) axis regulates cortisol levels and is particularly vulnerable to programming during foetal and neonatal development. Prenatal stress can disrupt HPA activity by reducing the efficiency of glucocorticoid feedback, leading to increased foetal brain exposure to cortisol. This increased exposure to stress is associated with decreased hippocampal volume and impaired cognitive abilities (12). Psychological, social, and physical stress during pregnancy can cause both acute and chronic spikes in cortisol levels, which are linked to higher concentrations of pro-inflammatory cytokines. These changes can negatively affect foetal growth and the gestational age at birth. (13). Notably, a study involving 86 mother-child dyads found that prenatal stress was associated with decreased hippocampal volume in newborns. This reduction in hippocampal volume was correlated with lower socio-emotional development in infants observed at 6 and 12 months of age (14). This evidence supports the hypothesis that the AOL experienced during pregnancy, particularly amid the COVID-19 pandemic, could adversely affect children’s neurological and socio-emotional development.

### 1.3 The impact of the COVID-19 pandemic on pregnancy outcomes, maternal mental health, and neurodevelopment

Pregnant women infected with SARS-CoV-2 generally exhibited moderate symptoms in 81-86% of cases, severe illness in 9.3-14%, and critical illness requiring hospitalization in an intensive care unit (ICU) in 5% of cases (15). COVID-19 during pregnancy increased the risk of severe neonatal morbidity by four times, and severe perinatal morbidity by 3.5 times, compared to pregnancies that tested negative for SARS-CoV-2 (16). Additionally, SARS-CoV-2 infection was linked to an increased risk of developing preeclampsia or eclampsia, which involves seizures in a hypertensive pregnancy (17). Vertical transmission, or transmission of the virus from a pregnant woman to her foetus in utero, was rare (18). However, maternal infection during the third trimester was associated with placental inflammation, increased Hofbauer cells (placental macrophages), heightened maternal immune response, thrombosis, placental infarcts, and poor fetoplacental perfusion (19,20). "Placentitis," or inflammation of the placenta, was observed in both severe and mild/asymptomatic cases of COVID-19 (21–23). During the COVID-19 pandemic, maternal depression rates varied significantly, ranging from 9.9% to 49% among participants. Anxiety symptoms were reported by 11% to 61% of participants, and 40% of women experienced stress symptoms (24). A cross-sectional study involving 7,645 participants found that the prevalence of depression and anxiety during pregnancy was 26.8% and 20.1%, respectively. In the postpartum period, these rates increased to 32.7% for depression and 26.6% for anxiety (25). In Brazil and Chile, during the pandemic, the prevalence of depression among pregnant women was 37.7% and 34.9%, respectively. Postpartum depression rates were even higher, with 47.2% in Brazil and 33.8% in Chile. Anxiety affected 34.3% of pregnant women in Brazil and 36% in Chile, with postpartum rates at 41.9% in Brazil and 41.5% in Chile (26). Regarding the birthing conditions during the pandemic, a study in the UK revealed that many pregnant women reported heightened anxiety and distress due to poor communication from hospital services, particularly concerning whether their birthing partners would be allowed to attend the birth (27). Similarly, in Chile, the restrictions placed on partners attending births during the first year of the pandemic probably caused a significant number of pregnant women to experience similar feelings of anxiety and distress, especially those who tested positive for COVID-19 (28).

Also, it is important to recognize that gender-based violence is a significant stressor during pregnancy, as intimate partner violence is linked to a higher prevalence of perinatal depression (29). Studies show that 37.0% of women who experienced physical intimate partner violence and 28.6% of women who faced sexual intimate partner violence reported depression (29). Additionally, during the lockdowns imposed due to the COVID-19 pandemic, emergency calls related to violence against women increased by 44% in Chile (26).

### 1.4 COVID-19 and the relationship between the mother and child

Prenatal stress during the COVID-19 pandemic has been linked to poorer socio-emotional development in infants (30). Additionally, higher levels of maternal postpartum depression have been associated with lower levels of attachment and bonding, increased parenting-related stress, and a reduced frequency of caregiving activities (31). Perinatal maternal depression may have negatively impacted the development of affectionate and emotional connections between mothers and their infants. The evidence indicates that postpartum depression during the COVID-19 pandemic predicted poorer social-emotional development in infants at six months of age (32). Furthermore, infection with SARS-CoV-2 during pregnancy increased the risk of neurodevelopmental disorders in one-year-old infants (OR 1.86)(33).

Parental stress (stress experienced by parents due to the demands and responsibilities of raising children) significantly increased in families that have experienced COVID-19, which positively correlated with self-perceived stress in children within the same family (34). Additionally, the parents who experienced economic and work problems during the COVID-19 pandemic reported a greater increase in negative parent-child interactions (35). Both parental stress and economic and work problems could be significant factors in the alterations of the mother-child relationship, early in the postnatal period or later during the parenting relationship after the COVID-19 pandemic.

## 2. Hypothesis

We hypothesise that the exposure of pregnant women to allostatic overload during the COVID-19 pandemic is independently associated with ischemic heart disease in mothers, neurodevelopmental alterations in children, and a dysfunctional relationship between the mother and child.

## 3. General objective

To assess the effect of allostatic overload during pregnancy in the context of the COVID-19 pandemic on the cardiovascular health of mothers, the neurodevelopment of children, and the relationship between the mother and child.

## 4. Specific Objectives

4.1 To assess the AOL experienced by women during pregnancy in the context of the COVID-19 pandemic.

4.1.1 To characterise the clinical evolution, mental health, social circumstances, and demographic factors of the mother during pregnancy.

4.1.2 To determine the prenatal AOL of the mother

4.2 To evaluate the cardiovascular health of mothers, the neurodevelopment of children, and the relationship between the mother and child.

4.2.1 To evaluate the cardiac function of mothers.

4.2.2 To evaluate the vascular function of mothers.

4.2.3 To quantify the cardiovascular plasma biomarkers in mothers.

4.2.4 To determine the neurodevelopmental status of children.

4.2.5 To appraise the mother-child relationship.

4.3 To assess the causal associations between exposure and outcomes and subsequent predictive models.

4.3.1 To evaluate the role of AOL during pregnancy and its association with ischemic heart disease in mothers in a controlled predictive model.

4.3.2 To evaluate the role of AOL during pregnancy and its association with neurodevelopmental disorders in children in a controlled predictive model.

4.3.3 To examine the role of AOL during pregnancy and its effect on the relationship between the mother and child, in a controlled predictive model.

## 5. Methods

### 5.1 Study design

This study will adopt a historical cohort design focusing on pregnant women during the COVID-19 pandemic (2020–2021). A mental health evaluation, conducted using the Edinburgh Postnatal Depression Scale (EPDS) applied during the prenatal period and a psychosocial evaluation to identify risk factors, will be registered from the clinical records. The AOL will be evaluated retrospectively through a clinimetric survey and a narrative interview conducted for a subsample, specifically focusing on the prenatal period to assess exposure.

For the exposure evaluation, we propose using a mixed-methods approach. This approach allows the investigator to collect and analyse data using both quantitative and qualitative methods. We can draw more comprehensive conclusions by integrating the findings from both methods (36). Specifically, we will employ simultaneous triangulation, which involves searching for corroboration and/or complementarity (or lack thereof) between quantitative and qualitative data related to AOL. Although the interaction between the two data groups during collection is limited, the findings will complement each other at the end of the study (36). Thus, the exposure evaluation will examine the correlation between the clinimetric survey results, and the data obtained from the prenatal EPDS application and psychosocial assessment, which will be extracted from clinical records. Meanwhile, the narrative interview, as a qualitative assessment, will provide a comprehensive understanding of the events and circumstances that may have led to the occurrence of AOL.

At present, we will evaluate significant parameters of the health of mothers, children, and the mother-child relationship. The primary outcomes of interest include the incidence of ischemic heart disease in mothers, neurodevelopmental alterations in children, and dysfunctional relationship between the mother and child. Figure 1 summarises the design of the study. This protocol study was evaluated and approved for funding by the National Agency of Research and Development (Agencia Nacional de Investigación y Desarrollo, ANID, Chile), Regular FONDECYT Competition, Project N°1241905.

**Figure 1.**
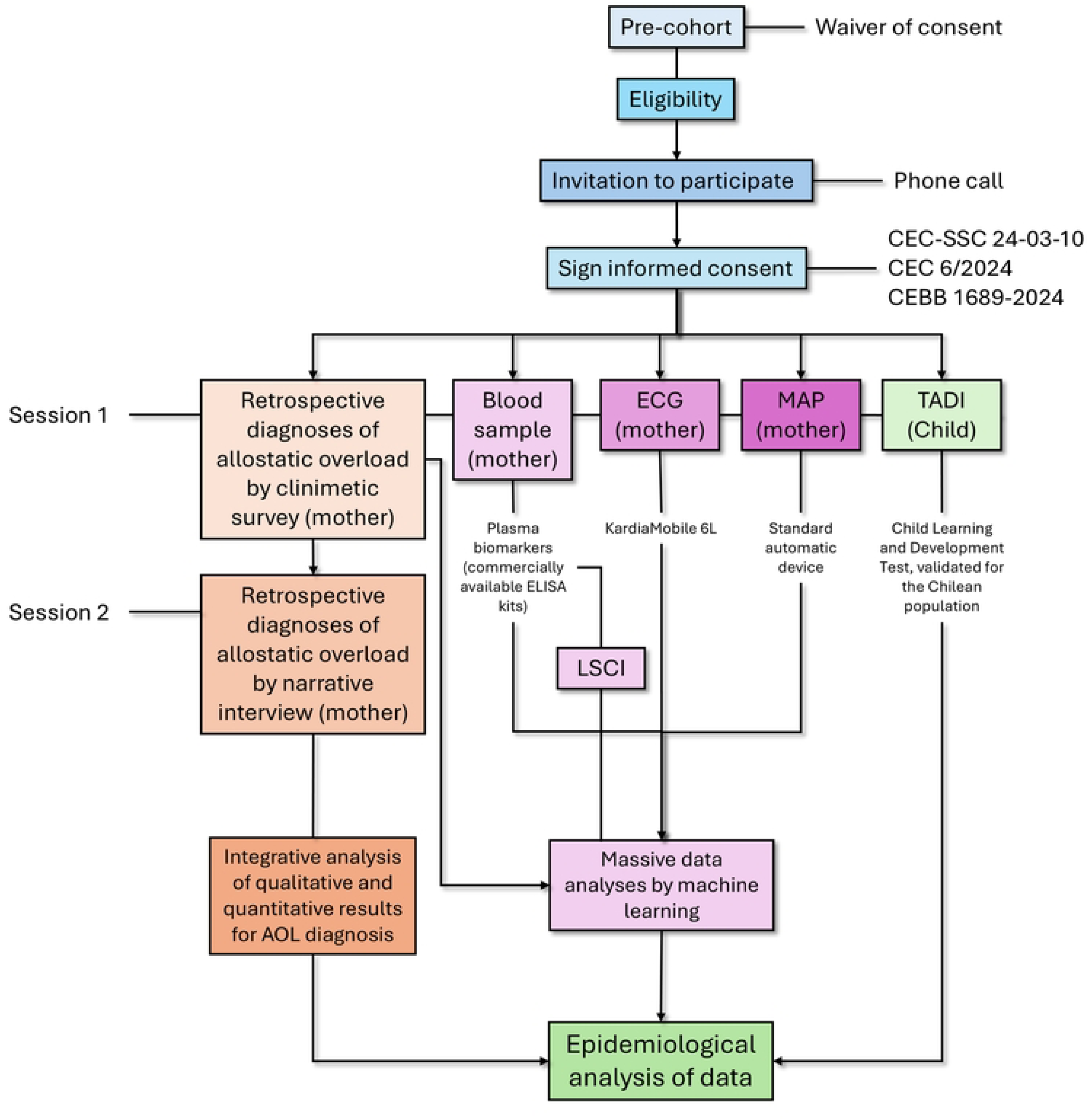
Design of the study. Historical cohort design, focusing on pregnant women during the COVID-19 pandemic (2020-2021). A pre-cohort will be established from clinical data (waiver of consent was approved by the Ethics and Scientific Committee of the Concepción Health Service), and eligibility will be determined according to inclusion and exclusion criteria. Mothers will be invited to participate via phone, and informed consent will be signed for her and their child (CEC-SSC 24-03-10, CEC 6/2024, CEBB 1689-2024). The clinimetric survey will be applied to the mother in session one, and the blood sample, ECG, and MAP will be taken. TADI will be applied to the children in a different room. In session two, the narrative interview (first 20 mothers) and later, the quantitative and qualitative data will be analysed to determine the diagnosis of allostatic overload. The blood sample will be frozen (-80^∘^C) until used to assess plasma biomarkers concentration by ELISA. Participants with significant alterations in endothelial dysfunction biomarkers will be invited to a follow-up session to assess the endothelium-dependent vasodilatation using Laser Speckle Contrast Imaging (LSCI). The quantitative data from the clinimetric survey, plasma biomarkers, ECG and MAP will be analysed using machine learning tools. Finally, the information on allostatic overload, machine learning results and TADI will be analysed using epidemiological methods.

### 5.2 Cohort formation

The cohort for this study will consist of mothers who carried out their pregnancies between March 1, 2020, and March 31, 2021, specifically during the first year of the COVID-19 pandemic (before vaccination of pregnant women). We will focus on pregnant women from Concepción (Bio-Bio Region, Chile) who received prenatal care during this period. Variables of interest, such as gender, sexual orientation, couple status, and previous pregnancies, will be considered potential third variables. Still, they will not be used as reasons for exclusion from the study. All mothers must sign an informed consent form to participate in the study.

The inclusion criteria for this study are as follows:

- Women who received prenatal care during the specified period and whose records are complete and legible for all variables of interest.

- Women whose children are currently alive.

- Women who have not given up their children for adoption.

- Singleton birth.

The exclusion criteria for this study are as follows:

- Women who had a pregnancy and birth during their adolescence.

- Women who do not have complete or adequate clinical records during pregnancy or did not receive prenatal care.

- Women who had pregnancies and births in other areas outside Concepción (Chile).

- Women for whom Spanish is not their first language.

- Women who give birth during incarceration, psychiatric hospitalisation, or under the supervision of child protection services (SENAME in Chile).

We are planning to conduct a study involving participants who are either exposed (with AOL) or unexposed (without AOL). We will include two unexposed participants as a comparison group for each exposed participant. Prior data indicate that the ischemic heart disease rate among controls is 0.001 (8). Suppose the true relative risk of failure for exposed subjects relative to controls is 1.83 (8). We must study 32 exposed and 64 unexposed participants to reject the null hypothesis that this relative risk equals 1 with a probability (power) of 0.8. The Type I error probability associated with this test of the null hypothesis is 0.05. We based this calculation on Fisher’s exact test for this null hypothesis.

#### 5.2.1 Recruitment of participants

A pre-cohort of participants was constituted from clinical records obtained from family health centres from Concepción (a waiver of consent was approved by the Ethics and Scientific Committee of the Concepción Health Service, CEC-SSC 24-03-10), considering women who had their pregnancies monitored between March 1, 2020, and March 31, 2021. At this stage, researchers had access to information that individually identified potential participants, which is necessary to later invite them to be part of the cohort. For the following stages, participating dyads will be assigned a code to anonymize the data stored in REDCap, and the information of those who do not agree to be part of the cohort will be removed from the study. The mothers will be invited to participate in the study via telephone, and those who agree will sign an informed consent form. This consent was previously revised and approved by the Ethics and Scientific Committee of the Concepción Health Service (CEC-SSC 24-03-10), Ethics Committee of the Faculty of Medicine of the Universidad de Concepción (CEC 6/2024), and Ethics, Bioethics and Biosafety Committee of the Vicerrectoría de Investigación y Desarrollo (VRID) of the Universidad de Concepción (CEBB 1689-2024). This consent includes the participation of their child.

### 5.3 Data collection

Study data will be collected and managed using REDCap electronic data capture tools hosted at the University of Concepción (37,38). REDCap (Research Electronic Data Capture) is a secure, web-based software platform designed to support data capture for research studies, providing 1) an intuitive interface for validated data capture; 2) audit trails for tracking data manipulation and export procedures; 3) automated export procedures for seamless data downloads to standard statistical packages; and 4) procedures for data integration and interoperability with external sources.

### 5.4 Allostatic overload determination

To determine the allostatic overload (AOL), we will use a mixed method, with data collected during prenatal care, a retrospective report through the clinimetric survey, and a narrative interview in a subset of participants.

#### 5.4.1 Clinimetric survey

The clinimetric survey to be collected is an adaptation of the evaluation proposed by Fava et al. (3). The survey asks participants to report on variables in the past tense for each pregnancy trimester, the period immediately before pregnancy, the postpartum period, and the time elapsed between the postpartum and the present day. Questions regarding COVID-19-related events (*e.g.*, contagion, deaths, isolation) have been added to the clinimetrics to capture contextual stressors. Table 1 lists the variables used for AOL determination.

**Table 1.** List of variables to exposure assessment: Allostatic Overload.

| Assessment of AOL |  | Retrospective clinimetric<br>(primary data collection) |
| --- | --- | --- |
| Prenatal care registries<br>(secondary data collection) | Health status and<br>sociodemographic<br>factors |  |
| Mental health risk<br>(Edinburgh Postnatal<br>Depression Scale). | COVID-19 contagion<br>(pregnancy period). | COVID-19 contagion of a<br>family member or close person. |
| Psychosocial risk<br>(Abbreviated Psychosocial<br>Assessment) | HIV detection during<br>pregnancy. | Death of a family member or<br>close person due to COVID-19. |
| Systolic blood pressure. | Marital status. | Death of a family member or<br>close person due to other<br>causes. |
| Diastolic blood pressure. | Venereal Disease<br>(VDRL). | Separation or divorce. |
| High-risk obstetric<br>pregnancy. | Level of education at<br>the moment of<br>pregnancy. | Change or loss of job. |
| Body mass index. | Absence of work<br>(pregnant woman<br>and/or partner). | Change of home. |
| Nutritional diagnosis. | Reduction or absence<br>of maternity leave. | Economic difficulties. |
| Gestational weight gain. | Drug abuse. | Legal problems. |
| Glycemia (26-28 weeks). | Smoking. | Pressure at work. |
| TTGO | Alcohol consumption | Pressure at home. |
| Placenta health. | Cesarean section. | Problems with coworkers. |
| Current pregnancy<br>planning. | Forceps or other birth<br>complications. | Problems with partner or other<br>family members. |
| Gender violence. |  | Victim of harassment. |
| Absence of companion at<br>controls |  | Illness of at least one family<br>member. |
| Absence of prenatal<br>controls. |  | Feeling that life was asking too<br>much of you. |
| Absence of birth<br>preparation. |  | Sleep disorders. |
| Pre-birth support (or pre-birth isolation). |  | General fatigue, lack of energy<br>(beyond typical pregnancy). |
| Birth support (or birth<br>isolation). |  | Feeling anxious, depressed,<br>irritable or sad. |
| Pre-term birth. |  | Demoralization. |
| Paternity lawsuits. |  | Caregiver for a person with<br>reduced mobility or an older<br>adult. |
| Alimony claims. |  | Caregiver for other children. |

#### 5.4.2 Narrative interviews

The first 20 women to accept to be part of the cohort will be invited to participate in the interviews using convenience sampling. Based on the results analysis, we will invite more participants if necessary. We selected this approach due to the accessibility of the study cohort. Although this approach may compromise the research process, the selection criteria applied to constitute the cohort ensure the quality of the qualitative data.

Generally, “a narrative interview takes the form of a conversation, and participants relate their experiences, bringing in whatever they consider relevant” (39). Narrative interviews provide the opportunity to prioritise the participant’s perspective, rather than relying on more specific information from previous findings, helping researchers better understand the experiences and behaviours. A narrative is not just a listing of events; it is a story in which the events have meaning and coherence, providing a context for a comprehensive understanding of the research phenomenon (40).

Rather than asking about pregnancy and its relationship to stressors, the interviewer will pose a broad thematic question: How did you experience your pregnancy? This way, the interviewee’s life experience will guide the search for information relevant to the research objectives. The interviews will respond to the collection of life experiences from the narratives of pregnant women during the COVID-19 pandemic. These will be framed by identifying and developing characteristics, context, crises, and significant events within a timeline with a clear beginning, middle, and end, focusing on the topic of interest.

The process for the data analysis consists of applying the phenomenological hermeneutical method of data analysis for multiple contexts developed by Morgan (41), which, in turn, is based on Lindseth and Norberg’s (42) proposal “phenomenological hermeneutical method for interpreting interview texts”. The procedure maintains the close and inevitable methodological relationship between phenomenology and hermeneutics as its foundation: “Thus we see that phenomenology must be phenomenological hermeneutics. Essential meaning must be studied and revealed in the interpretation of the text” (42).

Data analysis is structured around the stages of searching for meanings and contexts of interest, allowing for a phenomenological understanding of the text.

Naive reading: This first reading generates an intuitive and reflective approach to the text, setting aside the interpreter’s presuppositions. The text will be read several times to facilitate the researcher’s intuitive and reflective understanding.

Structural analysis: A theme conveys an essential meaning associated with a lived experience. As a result, to capture the meaning of what is lived within an experience, themes must be formulated not as abstract concepts but as concise descriptions to reveal their meaning. The whole text is read and divided into meaning units. A meaning unit can be part of a sentence, a sentence, several sentences, a paragraph, *i.e.* a piece of any length that conveys just one meaning (42). A set of meaning units gives meaning to a theme identified in the text. During structural analysis, naive reading will no longer be used. A comprehensive interpretation will be the cornerstone of understanding the texts in combined terms. The central idea highlights the contextual differences and similarities between the individual texts.

Comprehensive understanding: In this step, the main themes and sub-themes are summarised and reflected on in relation to the research question and the study context (42). To conclude the research process, it is necessary to conduct a comparative analysis of the analysed content in relation to its thematic significance and the state-of-the-art literature on the phenomenon of interest. This comparison enables the connection between the research participants’ life experiences and academic knowledge related to the phenomenon under investigation.

### 5.5 Cardiovascular health of the mothers

Cardiovascular health will be determined by mean arterial pressure (MAP), electrocardiography (ECG), heart rate (HR), and flow-mediated dilatation. Cardiovascular biomarkers will be studied to correlate with cardiovascular health parameters and other variables.

#### 5.5.6 Electrocardiography (ECG)

The ECG will be determined by the non-invasive KardiaMobile 6L device. The participant will place the device on their knee with their fingers, and a 30-second ECG will be recorded. The ECG will be printed and uploaded to the participant’s data. Each ECG will be taken in triplicate. ECG signals will undergo a secondary analysis. Using KardiaMobile 6L significantly reduces the time for ECG determination (43) and has shown feasibility and reliability compared to the standard 12-lead ECG (44).

The most significant ECG changes associated with cardiac disease are ST-segment changes, arrhythmias, and conduction disturbances (45). ECG diagnostic criteria for ischemic heart disease: The ST segment exhibits horizontal or hypotonic depression ≥0.1 mv (1.0 mm), sustained ≥1.0 min, and the interval between 2 episodes is ≥5.0 min. The typical ST segment downward measurement point is 80 ms after the J point and automatically becomes 50 ms after the J point when the heart rate is >120/min (46). The arrhythmias can be detected with R-R interval (time between heartbeats) and heart rate variability (HRV)—number of beats per minute (bpm). First-degree conduction block is detected by prolonging the P-R interval beyond 0.20 seconds (45).

#### 5.5.7 Assessment of cardiovascular dysfunction biomarkers

Fasting blood samples will be obtained by venipuncture and used to isolate plasma via centrifugation. All biomarkers will be determined by commercially available ELISA kits or validated methods. The list of biomarkers to determine are: soluble urokinase-type plasminogen activator receptor (suPAR)(47), growth differentiation factor 15 (GDF-15)(48), heart-type fatty acid-binding protein (H-FABP)(47), and soluble suppression of tumorigenicity 2 (sST2)(49) for coronary artery disease and ischemic heart failure; endothelin-1 (ET-1)(50), soluble vascular cell adhesion molecule-1 (sVCAM-1)(51), soluble tumour necrosis factor-α receptor I (sTNFRI)(52), asymmetric (ADMA), symmetric dimethylarginine (SDMA)(53), nitric oxide metabolites (54), and soluble fms-like tyrosine kinase-1 (sFlt-1)(55) for endothelial dysfunction; von Willebrand factor (vWF)(56), plasminogen activator inhibitor antigen (PAI)(57), and thrombomodulin (TM)(58) for coagulation/fibrinolysis:.

#### 5.5.8 Laser speckle contrast imaging (LSCI)

Participants with significant alterations in endothelial dysfunction biomarkers will be invited to a follow-up session to assess dermal blood flow (in the hand) using the Pericam® PSI-HR system (Perimed Ltd., Stockholm, Sweden)(59). LSCI can evaluate microcirculation in a non-contact, non-invasive manner with high spatial and temporal resolution (60). Post-occlusive reactive hyperaemia involves assessing the area under the curve of blood perfusion detected after the momentary release of previously occluded blood flow (61).

### 5.6 Children’s neurodevelopment and mother-child relationship

We will apply the Child Learning and Development Test, TADI (*Test de Aprendizaje y Desarrollo Infantil*)(TADI). TADI evaluates four dimensions: cognition, motor skills, language, and socio-emotionality; each constitutes an independent scale, where the items are ordered by increasing difficulty. It enables the evaluation of overall development, covering the four dimensions or each dimension separately, at three levels of development: Normal, Risk, and Delay (62). For maternal-child relationship assessment, we will use the Parenting Stress Index Short Form (PSI-SF), which is a self-report questionnaire designed to assess stress levels in four key domains: 1) Parental Distress, 2) Parent-Child Dysfunctional Interaction, and 3) Difficult Child. It comprises 36 items, most using a five-point Likert scale for responses (63).

### 5.7 Epidemiological statistical analysis

Statistical analyses will be performed using SPSS 18.0 Statistical Package Software for Windows (SPSS Inc., Chicago, IL, USA). The normality data of the variables will be evaluated with the Kolmogorov-Smirnov test. Continuous variables with normal distribution will be described using the mean and standard deviation; continuous variables without normal distribution will be described using the median and interquartile range. To analyse differences between independent groups, we will use the Student’s t-test (two-tailed) for normally distributed quantitative variables, the Mann-Whitney’s U-test for variables without normal distribution, and the Chi-square test for qualitative variables. Spearman’s correlation analyses will be used to evaluate correlations between different variables. All results will be considered statistically significant at p<0.05. For exposed and unexposed participants, incidence rates will be calculated for each outcome. We will perform a Cox Regression for the survival multivariable analysis to obtain adjusted hazard ratios (aHR). Time-to-event will be calculated in months, and the primary outcomes related to ischemic heart disease, neurodevelopmental disorders, and high parenting stress/defensive scores in the mother-child relationship assessment will be processed as binary variables indicating presence or absence. Models will be controlled for all relevant third variables that improve the model’s adjustment, consider confounders, reduce selection bias, and/or enhance precision.

### 5.8 Machine learning analysis

Exploratory analysis: Principal components analysis (PCA) will be performed using the software Pirouette® (Infometrix Inc., Bothell, WA, USA). Before pattern recognition, qualitative variables are transformed into categorical variables, and all data are pre-processed by autoscaling. PCA enables the determination of the data set’s structure and the identification of the latent factors responsible for its structure. Therefore, we will use PCA to identify the trends and persistence of significant clinical parameters related to the outcomes (64).

Classification analysis: Different subgroups of auto-scaled data will assess the prediction of ischemic heart disease in mothers, neurodevelopmental disorders in children, and relationship alterations between mothers and children through machine learning techniques: soft independent modelling of class analogy (SIMCA), partial least squares discriminant analysis (PLS-DA), and K Nearest Neighbour (KNN)(65).

Experimental evaluation of prediction models: Stratified approaches to N-Fold cross-validation. To begin with, the set of pre-classified patients will be divided into N equally sized (or almost similarly sized) groups, referred to as “folds”. In each of the N subsets, one is removed to be used only for testing (this guarantees that, in each run, a different testing set is used). The training is then carried out on the union of the remaining N-1 subsets (which contains approximately the exact representation of exposed and not exposed). The results will be averaged, and the standard deviation will be calculated.

Evaluation methods: Error rate (E) is the frequency of errors made by the prediction model over a given set of patients. It will be calculated by dividing the number of errors, N_FP_ (False positive) + N_FN_ (False negative), by the total number of patients, N_TP_ (True positive) + N_TN_ (True Negative) + N_FP_ + N_FN_. Accuracy (Acc) corresponds to the frequency of correct classifications made by the prediction model over a given set of patients. Classification accuracy will be calculated by dividing the number of correct classifications, N_TP_ + N_TN_, by the total number of examples. Precision (Pre) corresponds to the percentage of true positives, N_TP_, among all patients that the prediction model has labelled as positive: N_TP_ + N_FP_. Sensitivity (Se) corresponds to the probability that a positive patient will be correctly recognised as such (by the prediction model). The value will be, therefore, obtained by dividing the number of true positives, N_TP_, by the number of positives in the given set: N_TP_ + N_FN_. Specificity (Sp) corresponds to the probability that a negative patient will be correctly recognised as such. The value will be, therefore, obtained by dividing the number of true negatives, N_TN_, by the number of positives in the given set: N_TN_ + N_FP_. Gmean corresponds to the geometric mean, telling us what Se and Sp are different. It will be calculated by the square root of the product of sensitivity and specificity.

### 5.9 Timeline

The pre-cohort was formed from clinical records (waiver of informed consent authorized) between October 7, 2024, and January 31, 2025. The pilot tests of the protocol were conducted from March to April 2025 to finalize the methods, clinimetric survey, and the REDCap instrument by the end of April 2025. Participant recruitment for the cohort will begin on June 2, 2025, and will continue until July 31, 2026. Data collection will be completed alongside the recruitment process. A comprehensive analysis of the data and the main results of the study are expected to be obtained by December 2026.

## 6. Discussion

The COVID-19 pandemic has exposed pregnant women to an unprecedented level of stress, resulting in higher levels of prenatal anxiety, depression, and stress. The impact of COVID-19 on maternal mortality and maternal and neonatal morbidity is well-documented (16,66–70). The impact is not exclusively related to the biological effects of SARS-CoV-2 infection but is also associated with accumulating stressors that negatively affect the health of mothers and neonates (16,25,33). The complexity of the pandemic period for pregnant women invites us to build a comprehensive conceptual framework that integrates knowledge of stress physiology, neurobiology, and pregnancy physiology (71). Including the concept of AOL as an exposure variable enables us to take a step towards constructing this comprehensive model. It allows us to consider factors beyond the current gaps in knowledge of maternal health and merge both the adaptive physiological processes of pregnancy, labour, and birth, as well as the increased susceptibility to long-term pathologies that require diagnosis, prevention, and treatment. The expected results of this study will be useful in determining if the AOL evaluation during pregnancy could be a good tool for understanding the physical and mental health consequences of dealing with prolonged stress (3,72). Including AOL in clinical records may serve as an essential health indicator, especially for pregnant women living in more vulnerable conditions, who are at high risk of suffering stress and AOL.

AOL is a holistic concept that incorporates a variety of harmful elements that were exposed to pregnant mothers during the pandemic, affecting their physiological adaptations during pregnancy. As this study protocol proposes to determine, the complexity of AOL is also related to the health outcomes affected, including the mother’s cardiovascular health, the neurodevelopment of children, and the maternal-child relationship. This study will use quantitative and qualitative methods to determine the exposure, combined with sophisticated statistical analysis, including machine learning tools, to comprehensively understand the phenomenon experienced by pregnant women during the COVID-19 pandemic and its consequences.

In this protocol study, the AOL will be determined by examining the clinical records that were previously collected during pregnancy. These records include AOL markers like mean arterial pressure, gestational weight gain, glycemia, mental health and psychosocial risk, reduction or absence of maternity leave, gender violence, economic difficulties, isolation in prenatal controls or birth, lack of maternal-newborn bonding, prenatal anxiety and depression, among others, all variables that could have been significantly altered by the COVID-19 pandemic conditions. Mothers may have different capacities for coping with these difficulties, and this protocol was designed to classify and evaluate the consequences of inadequate response to the challenges for the mother-child dyads.

There is evidence that AOL increases cardiovascular risk (5), and perinatal AOL is associated with preterm birth (73), while prenatal depression increases the prevalence of chronic hypertension later in life in women (8). Due to the evidence of increased perinatal mental health disorders during the COVID-19 pandemic in Latin America and Chile (26), we will consider the perinatal anxiety, depression and stress, and factors that could be cause or mediate mental health disorders, like economic difficulties, lack of social support, and gender violence, as relevant factors to determine the presence of AOL during the pregnancy.

Regarding mental health in prenatal controls, the Edinburgh Postnatal Depression Scale (EPDS) is a 10-item scale for the identification of maternal depression, applied in the Chilean public system in the prenatal and postnatal periods. Each item contains short descriptive statements, scoring the most negative description with the highest value of 3 and the most positive with 0. Scores are tallied so that a score <8 indicates depression is not likely, 9–11 indicates depression is possible, 12–13 indicates a reasonably high possibility of depression, and 14 and higher is considered a positive screen for depression and recommends a diagnostic assessment. Additionally, a positive score is identified for question 10, which identifies participants at risk of harm or suicide (74,75). The Abbreviated Psychosocial Assessment (EPsA) is an instrument for early detection and intervention of some risk factors that introduce inequity in people’s development. This evaluation is considered a priority strategy for monitoring and supporting pregnancy in the Chilean pregnancy and early childhood protection system, "Chile Crece Contigo (ChCC)" (76). These data, more other information will be collected in the clinimetric survey as is shown in table 1. Following the original protocol from Fava (3) and modifications proposed by Peng (77), it is possible to determine a score of AL for each participant. The possibility of determining a score of AL and evaluating the life experience associated with AOL to determine exposure is a strategy to overcome the weaknesses of quantitative or qualitative methods themselves (36). By applying data triangulation, we can gain diverse perspectives on the COVID-19 pandemic and achieve a deeper understanding of this phenomenon. This approach allows us to effectively distinguish between women who experienced allostatic overload and those who managed to cope adequately with the challenges presented during the pandemic.

One advantage of this study design is the ability to assess the long-term effects of COVID-19 during pregnancy. Long COVID, also known as post-COVID-19 syndrome, is estimated to affect tens of millions of people. However, there is still a lack of information regarding the mechanisms or biomarkers that could aid in the classification and treatment of this condition (78). Among the potential consequences of long COVID, cardiovascular diseases have surfaced as a significant concern for the healthcare system, particularly for patients who already have pre-existing conditions such as obesity, diabetes, or other risk factors for cardiovascular disease (79). Given the characteristics of Chilean pregnant women—approximately 30% are obese (80), and around 10% experience gestational diabetes in the Biobío region (81)—there is a possibility that we will observe a percentage of participants that experienced symptoms of long COVID, especially those associated with cardiovascular disease. This will be an additional finding of our study, which will contribute to the understanding of cardiovascular biomarkers related to long COVID (82), particularly in a unique population like women who had a pregnancy with SARS-CoV-2 infection.

Finally, we will use machine learning (ML) to analyse the data besides classic statistical tools. ML corresponds to various methods that use mathematics, statistics, and computational science to learn from multivariate data. Employing pattern recognition performed on various measured variables, different algorithms can find correlations and accurately predict different conditions, such as an individual belonging to a particular group or class or the concentration of a specific biomarker in a sample of interest (65,83). ML approaches are available for managing large and heterogeneous data sources, identifying intricate and occult patterns, and predicting complex outcomes (64). The pregnancy window of time represents an excellent opportunity to exploit all the advantages of using ML, especially for accurately predicting short- and long-term adverse outcomes.

## 7. Strengths and weaknesses of the protocol study

One strength of conducting a retrospective evaluation of exposure is the access to a substantial amount of information from pregnancy control data, particularly in the unique context of the COVID-19 pandemic. This approach also allows for the evaluation of mother-child dyads who were not selected to be part of a study during the exposure period, so they experienced the challenges of the pandemic as part of the "real world." Collecting information from pregnancy controls within the public health system offers the advantage of standardized formats and consistent clinical records, which help minimize potential biases associated with data collection. Retrospective studies are particularly useful for examining specific periods, such as the COVID-19 pandemic, and identifying risk factors related to diseases or clinical conditions long after exposure. This study aims to evaluate outcomes today, 4 to 5 years after exposure to pandemic conditions. This timeframe is adequate for assessing the long-term effects of pregnancies developed in stressful situations, and the implications for the mother and child.

On the other hand, a significant weakness of the retrospective AOL evaluation is recall bias. Mothers may have difficulty remembering specific experiences or selectively recall those they feel are more appropriate to share during the interviews. Additionally, individuals might exaggerate or downplay the severity of stressors to present a more favourable narrative to the interviewer. When assessing the risk of bias in the narrative interviews, it is important to consider their current emotional state or feelings related to the stressors.

To address these potential weaknesses, we will conduct interviews with multiple participants. After analysing the initial interviews, we may increase the number of interviews if we encounter controversial or unclear information. Furthermore, we will compare the information gathered from the interviews with data from clinical records, which provide verifiable information obtained during the exposure period.

## 8. Expected results

From quantitative results:

- Higher detection of AOL in women who had pregnancy during the first wave of the COVID-19 pandemic, compared with the after-period.
- Higher risk of endothelial dysfunction in women exposed to AOL.
- Higher risk of ECG alterations (ischemic heart disease, arrhythmia) and HRV in women exposed to AOL.
- Higher risk of neurodevelopmental disorders in children born from AOL pregnancies.
- Alterations in mother-child relationships when the mother was exposed to AOL.
- A predicted model for cardiovascular risk in women after pregnancy with AOL. From qualitative results:
- A comprehensive understanding of AOL phenomena in a Latino context from the day- to-day life experiences of mothers during the pandemic.
- To identify other stress factors not evaluated by quantitative methods.

## Data Availability

No datasets were generated or analysed during the current study. All relevant data from this study will be made available upon study completion.

## References

1. Fava GA, Cosci F, Sonino N. Current Psychosomatic Practice. Psychother Psychosom. 2017;86(1):13–30.

2. McEwen BS, Stellar E. Stress and the individual. Mechanisms leading to disease. Arch Intern Med. 27 de septiembre de 1993;153(18):2093-101.

3. Fava GA, McEwen BS, Guidi J, Gostoli S, Offidani E, Sonino N. Clinical characterization of allostatic overload. Psychoneuroendocrinology. octubre de 2019;108:94–101.

4. Doan SN. Allostatic load: Developmental and conceptual considerations in a multi-system physiological indicator of chronic stress exposure. Dev Psychobiol. 2021;63(5):825–36.

5. Guidi J, Lucente M, Piolanti A, Roncuzzi R, Rafanelli C, Sonino N. Allostatic overload in patients with essential hypertension. Psychoneuroendocrinology. 1 de marzo de 2020;113:104545.

6. Hux VJ, Roberts JM, Okun ML. Allostatic load in early pregnancy is associated with poor sleep quality. Sleep Med. 1 de mayo de 2017;33:85-90.

7. Barrett ES, Vitek W, Mbowe O, Thurston SW, Legro RS, Alvero R, et al. Allostatic load, a measure of chronic physiological stress, is associated with pregnancy outcomes, but not fertility, among women with unexplained infertility. Hum Reprod. 1 de septiembre de 2018;33(9):1757-66.

8. Ackerman-Banks CM, Lipkind HS, Palmsten K, Pfeiffer M, Gelsinger C, Ahrens KA. Association of Prenatal Depression With New Cardiovascular Disease Within 24 Months Postpartum. J Am Heart Assoc. 2 de mayo de 2023;12(9):e028133.

9. Khosla K, Heimberger S, Nieman KM, Tung A, Shahul S, Staff AC, et al. Long-Term Cardiovascular Disease Risk in Women After Hypertensive Disorders of Pregnancy: Recent Advances in Hypertension. Hypertension. octubre de 2021;78(4):927–35.

10. Cirillo PM, Cohn BA. Pregnancy Complications and Cardiovascular Disease Death. Circulation. 29 de septiembre de 2015;132(13):1234-42.

11. Barker DJP. The origins of the developmental origins theory. J Intern Med. mayo de 2007;261(5):412–7.

12. Matthews SG. Early programming of the hypothalamo-pituitary-adrenal axis. Trends Endocrinol Metab TEM. noviembre de 2002;13(9):373–80.

13. Hunter PJ, Awoyemi T, Ayede AI, Chico RM, David AL, Dewey KG, et al. Biological and pathological mechanisms leading to the birth of a small vulnerable newborn. The Lancet. 20 de mayo de 2023;401(10389):1720-32.

14. Moog NK, Nolvi S, Kleih TS, Styner M, Gilmore JH, Rasmussen JM, et al. Prospective association of maternal psychosocial stress in pregnancy with newborn hippocampal volume and implications for infant social-emotional development. Neurobiol Stress. 1 de noviembre de 2021;15:100368.

15. Ayala-Ramírez P, González M, Escudero C, Quintero-Arciniegas L, Giachini FR, Alves de Freitas R, et al. Severe Acute Respiratory Syndrome Coronavirus 2 Infection in Pregnancy. A Non-systematic Review of Clinical Presentation, Potential Effects of Physiological Adaptations in Pregnancy, and Placental Vascular Alterations. Front Physiol. 2022;13:785274.

16. Villar J, Ariff S, Gunier RB, Thiruvengadam R, Rauch S, Kholin A, et al. Maternal and Neonatal Morbidity and Mortality Among Pregnant Women With and Without COVID-19 Infection: The INTERCOVID Multinational Cohort Study. JAMA Pediatr. 22 de abril de 2021;

17. Ferrara A, Hedderson MM, Zhu Y, Avalos LA, Kuzniewicz MW, Myers LC, et al. Perinatal Complications in Individuals in California With or Without SARS-CoV-2 Infection During Pregnancy. JAMA Intern Med. 1 de mayo de 2022;182(5):503-12.

18. Edlow AG, Li JZ, Collier ARY, Atyeo C, James KE, Boatin AA, et al. Assessment of Maternal and Neonatal SARS-CoV-2 Viral Load, Transplacental Antibody Transfer, and Placental Pathology in Pregnancies During the COVID-19 Pandemic. JAMA Netw Open. 1 de diciembre de 2020;3(12):e2030455.

19. González M, Troncoso F, Escudero C, González M, Troncoso F, Escudero C. SARS-CoV- 2 (COVID-19) en gestación y placenta: una revisión narrativa sobre el estado del arte. Rev Chil Obstet Ginecol. agosto de 2021;86(4):425–32.

20. Schwartz DA, Mulkey SB, Roberts DJ. SARS-CoV-2 placentitis, stillbirth, and maternal COVID-19 vaccination: clinical–pathologic correlations. Am J Obstet Gynecol. 1 de marzo de 2023;228(3):261-9.

21. Sharps MC, Hayes DJL, Lee S, Zou Z, Brady CA, Almoghrabi Y, et al. A structured review of placental morphology and histopathological lesions associated with SARS-CoV-2 infection. Placenta. noviembre de 2020;101:13–29.

22. Stenton S, McPartland J, Shukla R, Turner K, Marton T, Hargitai B, et al. SARS-COV2 placentitis and pregnancy outcome: A multicentre experience during the Alpha and early Delta waves of coronavirus pandemic in England. EClinicalMedicine. mayo de 2022;47:101389.

23. Ward JD, Cornaby C, Kato T, Gilmore RC, Bunch D, Miller MB, et al. The clinical impact of maternal COVID-19 on mothers, their infants, and placentas with an analysis of vertical transfer of maternal SARS-CoV-2-specific IgG antibodies. Placenta. 1 de junio de 2022;123:12-23.

24. Wall S, Dempsey M. The effect of COVID-19 lockdowns on women’s perinatal mental health: a systematic review. Women Birth. 1 de febrero de 2023;36(1):47-55.

25. Mesquita A, Costa R, Bina R, Cadarso-Suárez C, Gude F, Díaz-Louzao C, et al. A cross- country study on the impact of governmental responses to the COVID-19 pandemic on perinatal mental health. Sci Rep. 16 de febrero de 2023;13(1):2805.

26. González-Ortiz M, Castro P, Vergara P, Huerta P, Escudero C. COVID-19 on Pregnancy Outcomes, Mental Health and Placenta: Focus in Latin America. En: Advances in Maternal-Fetal Biomedicine: Cellular and Molecular Mechanisms of Pregnancy Pathologies. Switzerland: Springer Nature; 2023. (Advances in Experimental Medicine and Biology).

27. Aydin E, Glasgow KA, Weiss SM, Khan Z, Austin T, Johnson MH, et al. Giving birth in a pandemic: women’s birth experiences in England during COVID-19. BMC Pregnancy Childbirth. 2022;22(1).

28. Leiva G, Sadler M, Quezada S, Flores V, Sierra C, Díaz S, et al. Respuesta al COVID en una maternidad chilena.pdf. 2021.

29. Roddy Mitchell A, Gordon H, Lindquist A, Walker SP, Homer CSE, Middleton A, et al. Prevalence of Perinatal Depression in Low- and Middle-Income Countries: A Systematic Review and Meta-analysis. JAMA Psychiatry [Internet]. 8 de marzo de 2023 [citado 10 de marzo de 2023]; Disponible en: 10.1001/jamapsychiatry.2023.0069

30. Duguay G, Garon-Bissonnette J, Lemieux R, Dubois-Comtois K, Mayrand K, Berthelot N. Socioemotional development in infants of pregnant women during the COVID-19 pandemic: the role of prenatal and postnatal maternal distress. Child Adolesc Psychiatry Ment Health. 2022;16(1).

31. Federica G, Renata T, Marzilli E. Parental Postnatal Depression in the Time of the COVID-19 Pandemic: A Systematic Review of Its Effects on the Parent–Child Relationship and the Child’s Developmental Outcomes. Int J Environ Res Public Health. enero de 2023;20(3):2018.

32. Harrison M, Rohde J, Hatchimonji D, Berman T, Flatley CA, Okonak K, et al. Postpartum Mental Health and Infant Development During the COVID-19 Pandemic. Pediatrics. 23 de febrero de 2022;149(1 Meeting Abstracts February 2022):87.

33. Edlow AG, Castro VM, Shook LL, Kaimal AJ, Perlis RH. Neurodevelopmental Outcomes at 1 Year in Infants of Mothers Who Tested Positive for SARS-CoV-2 During Pregnancy. JAMA Netw Open. 9 de junio de 2022;5(6):e2215787.

34. Di Chiara C, Ferrarese M, Boracchini R, Cantarutti A, Tibaldo AL, Stefanni C, et al. Long-term neuropsychiatric and neuropsychological impact of the pandemic in Italian COVID-19 family clusters, including children and parents. PloS One. 2025;20(4):e0321366.

35. Oftedal A, Larsen L, Helland MS. Economic Hardship During the Covid-19 Pandemic and Trajectories of Parent–Child Relationships: A Prospective Longitudinal Study Among Norwegian Families. Fam Process. 2025;64(2):e70031.

36. Arias Valencia MM. Principles, Scope, and Limitations of the Methodological Triangulation. Investig Educ En Enfermeria. 15 de septiembre de 2022;40(2):e03.

37. Harris PA, Taylor R, Minor BL, Elliott V, Fernandez M, O’Neal L, et al. The REDCap consortium: Building an international community of software platform partners. J Biomed Inform. 1 de julio de 2019;95:103208.

38. Harris PA, Taylor R, Thielke R, Payne J, Gonzalez N, Conde JG. Research electronic data capture (REDCap)—A metadata-driven methodology and workflow process for providing translational research informatics support. J Biomed Inform. 1 de abril de 2009;42(2):377-81.

39. Bates JA. Use of narrative interviewing in everyday information behavior research. Libr Inf Sci Res. 1 de diciembre de 2004;26(1):15-28.

40. Anderson C, Kirkpatrick S. Narrative interviewing. Int J Clin Pharm. 1 de junio de 2016;38(3):631-4.

41. Morgan DA. Analysing complexity: developing a modified phenomenological hermeneutical method of data analysis for multiple contexts. Int J Soc Res Methodol. 2 de noviembre de 2021;24(6):655-67.

42. Lindseth A, Norberg A. A phenomenological hermeneutical method for researching lived experience. Scand J Caring Sci. junio de 2004;18(2):145–53.

43. Gaddam M, Liu A, Lohrmann G, Breed A, Passman R. KardiaMobile 6L versus 12-lead ECG: Effects on clinic utilization time. J Cardiovasc Electrophysiol. 2024;35(8):1691–3.

44. Zaballos M, Fernández I, Rodríguez L, Orozco S, García A, Juncos M, et al. Feasibility of using KardiaMobile-L6 for QT interval monitoring during the early phase of the COVID-19 pandemic in critical care patients. Sci Rep. 6 de julio de 2023;13(1):10985.

45. Irwin S. Clinical Manifestations and Assessment of Ischemic Heart Disease. Phys Ther. 1 de diciembre de 1985;65(12):1806-11.

46. Liu Y, Ping J, Qiu L, Sun C, Chen M. Comparative analysis of ischemic changes in electrocardiogram and coronary angiography results. Medicine (Baltimore). 18 de junio de 2021;100(24):e26007.

47. Fröhling T, Semo D, Mirna M, Paar V, Shomanova Z, Motloch LJ, et al. Novel Biomarkers as Potential Predictors of Decompensated Advanced Chronic Heart Failure-Single Center Study. J Clin Med. 14 de noviembre de 2024;13(22):6866.

48. Dalos D, Spinka G, Schneider M, Wernly B, Paar V, Hoppe U, et al. New Cardiovascular Biomarkers in Ischemic Heart Disease—GDF-15, A Probable Predictor for Ejection Fraction. J Clin Med. 27 de junio de 2019;8(7):924.

49. Maisel AS, Di Somma S. Do we need another heart failure biomarker: focus on soluble suppression of tumorigenicity 2 (sST2). Eur Heart J. 7 de agosto de 2017;38(30):2325-33.

50. Willems LH, Nagy M, ten Cate H, Spronk HMH, Groh LA, Leentjens J, et al. Sustained inflammation, coagulation activation and elevated endothelin-1 levels without macrovascular dysfunction at 3 months after COVID-19. Thromb Res. 2022;209:106-14.

51. Geraets AFJ, van Agtmaal MJM, Stehouwer CDA, Sörensen BM, Berendschot TTJM, Webers CAB, et al. Association of Markers of Microvascular Dysfunction With Prevalent and Incident Depressive Symptoms: The Maastricht Study. Hypertens Dallas Tex 1979. agosto de 2020;76(2):342-9.

52. Palomo M, Youssef L, Ramos A, Torramade-Moix S, Moreno-Castaño AB, Martinez-Sanchez J, et al. Differences and similarities in endothelial and angiogenic profiles of preeclampsia and COVID-19 in pregnancy. Am J Obstet Gynecol. 26 de marzo de 2022;S0002-9378(22)00227-7.

53. Bima C, Parasiliti-Caprino M, Rumbolo F, Ponzetto F, Gesmundo I, Nonnato A, et al. Asymmetric and symmetric dimethylarginine as markers of endothelial dysfunction in cerebrovascular disease: A prospective study. Nutr Metab Cardiovasc Dis NMCD. julio de 2024;34(7):1639–48.

54. Bryan NS, Grisham MB. Methods to Detect Nitric Oxide and its Metabolites in Biological Samples. Free Radic Biol Med. 1 de septiembre de 2007;43(5):645-57.

55. Abozeid SF, Elkholy RA, Elbedewy TA, Seliem MF. Soluble Fms-like tyrosine kinase-1 as an endothelial dysfunction biomarker associated with pulmonary hypertension in adult patients with beta-thalassemia major. J Investig Med Off Publ Am Fed Clin Res. diciembre de 2024;72(8):883–90.

56. Tavares IR, Caffaro RA, Portugal MF, Ribeiro CM, da Silva VS, Krupa E, et al. Biomarkers Profile in Provoked Versus Unprovoked Deep Venous Thrombosis. Clin Appl Thromb Off J Int Acad Clin Appl Thromb. 2024;30:10760296241238211.

57. Yatsenko T, Rios R, Nogueira T, Takahashi S, Tabe Y, Naito T, et al. Urokinase-type plasminogen activator and plasminogen activator inhibitor-1 complex as a serum biomarker for COVID-19. Front Immunol. 2023;14:1299792.

58. Padilla S, Andreo M, Marco P, Marco-Rico A, Ledesma C, Fernández-González M, et al. Enhanced prediction of thrombotic events in hospitalized COVID-19 patients with soluble thrombomodulin. PloS One. 2025;20(3):e0319666.

59. Meriño M, Martín SS, Sandaña P, Herlitz K, Aguayo C, Godoy A, et al. Deletion of the adenosine A2A receptor increases the survival rate in a mice model of polymicrobial sepsis. Purinergic Signal. 1 de septiembre de 2020;16(3):427-37.

60. Vág J, Nagy TL, Mikecs B. Sex-related differences in endothelium-dependent vasodilation of human gingiva. BMC Oral Health. 13 de mayo de 2022;22(1):177.

61. Roustit M, Cracowski JL. Assessment of endothelial and neurovascular function in human skin microcirculation. Trends Pharmacol Sci. julio de 2013;34(7):373–84.

62. Schonhaut L, Edwards M, Pardo M, Valdés A. [Properties of the Test of Early Learning and Development, second edition «TADI», in the context of scale validation policies for children under 6 years of age in Chile and Latin America]. Andes Pediatr Rev Chil Pediatr. agosto de 2024;95(4):353–63.

63. Abidin R, Flens JR, Austin WG. The Parenting Stress Index. En: Forensic uses of clinical assessment instruments. Mahwah, NJ, US: Lawrence Erlbaum Associates Publishers; 2006. p. 297-328.

64. Araya J, Rodriguez A, Lagos-SanMartin K, Mennickent D, Gutiérrez-Vega S, Ortega-Contreras B, et al. Maternal thyroid profile in first and second trimester of pregnancy is correlated with gestational diabetes mellitus through machine learning. Placenta. 1 de enero de 2021;103:82-5.

65. Mennickent D, Ortega-Contreras B, Gutiérrez-Vega S, Castro E, Rodríguez A, Araya J, et al. Evaluation of first and second trimester maternal thyroid profile on the prediction of gestational diabetes mellitus and post load glycemia. PloS One. 2023;18(1):e0280513.

66. Hessami K, Homayoon N, Hashemi A, Vafaei H, Kasraeian M, Asadi N. COVID-19 and maternal, fetal and neonatal mortality: a systematic review. J Matern-Fetal Neonatal Med Off J Eur Assoc Perinat Med Fed Asia Ocean Perinat Soc Int Soc Perinat Obstet. 16 de agosto de 2020;1-6.

67. Lumbreras-Marquez MI, Campos-Zamora M, Leon HLD de, Farber MK. Maternal mortality from COVID-19 in Mexico. Int J Gynecol Obstet. 2020;150(2):266–7.

68. 68. Maza-Arnedo F, Paternina-Caicedo A, Sosa CG, Mucio B de, Rojas-Suarez J, Say L, et al. Maternal mortality linked to COVID-19 in Latin America: Results from a multi-country collaborative database of 447 deaths. Lancet Reg Health – Am [Internet]. 1 de agosto de 2022 [citado 8 de marzo de 2023];12. Disponible en: https://www.thelancet.com/journals/lanam/article/PIIS2667-193X(22)00086-2/fulltext#seccesectitle0017

69. Mendez-Dominguez N, Santos-Zaldívar K, Gomez-Carro S, Datta-Banik S, Carrillo G. Maternal mortality during the COVID-19 pandemic in Mexico: a preliminary analysis during the first year. BMC Public Health. 2 de julio de 2021;21(1):1297.

70. Thoma ME, Declercq ER. All-Cause Maternal Mortality in the US Before vs During the COVID-19 Pandemic. JAMA Netw Open. 28 de junio de 2022;5(6):e2219133.

71. 71. Dickens MJ, Pawluski JL, Romero LM. Moving Forward From COVID-19: Bridging Knowledge Gaps in Maternal Health With a New Conceptual Model. Front Glob Womens Health [Internet]. 2020 [citado 8 de junio de 2023];1. Disponible en: https://www.frontiersin.org/articles/10.3389/fgwh.2020.586697

72. Guidi J, Lucente M, Sonino N, Fava GA. Allostatic Load and Its Impact on Health: A Systematic Review. Psychother Psychosom. 2021;90(1):11–27.

73. 73. Premji SS, Pana GS, Cuncannon A, Ronksley PE, Dosani A, Hayden KA, et al. Prenatal allostatic load and preterm birth: A systematic review. Front Psychol [Internet]. 2022 [citado 13 de junio de 2023];13. Disponible en: https://www.frontiersin.org/articles/10.3389/fpsyg.2022.1004073

74. Jadresic E, Araya R, Jara C. Validation of the Edinburgh Postnatal Depression Scale (EPDS) in Chilean postpartum women. J Psychosom Obstet Gynaecol. diciembre de 1995;16(4):187–91.

75. Cox JL, Holden JM, Sagovsky R. Detection of postnatal depression. Development of the 10-item Edinburgh Postnatal Depression Scale. Br J Psychiatry J Ment Sci. junio de 1987;150:782-6.

76. Chile Crece Contigo. Riesgos psicosociales durante la gestación: violencia de género [Internet]. 2011 [citado 30 de junio de 2023]. Disponible en: http://www.chccsalud.cl/2011/12/riesgos-psicosociales-durante-la.html

77. Peng M, Wang L, Xue Q, Yin L, Zhu BH, Wang K, et al. Post-COVID-19 Epidemic: Allostatic Load among Medical and Nonmedical Workers in China. Psychother Psychosom. 2021;90(2):127–36.

78. Dieter RS, Kempaiah P, Dieter EG, Alcazar A, Tafur A, Gerotziafas G, et al. Cardiovascular Symposium on Perspectives in Long COVID. Clin Appl Thromb. 1 de abril de 2025;31:10760296251319963.

79. Tsampasian V, Bäck M, Bernardi M, Cavarretta E, Dębski M, Gati S, et al. Cardiovascular disease as part of Long COVID: a systematic review. Eur J Prev Cardiol. 1 de abril de 2025;32(6):485-98.

80. Bertini A, Varela MJ, Holz A, Gonzalez P, Bastias D, Giovanetti M, et al. Impact of pregestational obesity on perinatal complications: update in a Latin American cohort. Public Health. 1 de agosto de 2024;233:170-6.

81. González-Ortiz M, Cisterna M, Henning R, Cisterna C, Castro P, Escudero C, et al. Incidencia de diabetes gestacional en Chile durante el periodo 2001-2022. Rev Chil Obstet Ginecol. 24 de mayo de 2024;89(2):100-8.

82. 82. Lai YJ, Liu SH, Manachevakul S, Lee TA, Kuo CT, Bello D. Biomarkers in long COVID-19: A systematic review. Front Med [Internet]. 20 de enero de 2023 [citado 13 de mayo de 2025];10. Disponible en: https://www.frontiersin.org/journals/medicine/articles/10.3389/fmed.2023.1085988/full

83. 83. Mennickent D, Rodríguez A, Opazo MaC, Riedel CA, Castro E, Eriz-Salinas A, et al. Machine learning applied in maternal and fetal health: a narrative review focused on pregnancy diseases and complications. Front Endocrinol [Internet]. 2023 [citado 15 de junio de 2023];14. Disponible en: https://www.frontiersin.org/articles/10.3389/fendo.2023.1130139

